# Applying Deep Learning in Heart Failure: Hospital Readmission is Not Like Other Health Quality Metrics

**DOI:** 10.1101/2024.03.27.24304999

**Authors:** Hailey M. Shepherd, Jeffrey T. Heaton, Theodore Marghitu, Yun Z. Bai, Melanie P. Subramanian, Sophia H. Roberts, Martha M.O. McGilvray, Amit A. Pawale, Gregory A. Ewald, Brian P. Cupps, Michael K. Pasque, Randi E. Foraker

**Affiliations:** Division of Cardiothoracic Surgery, Department of Surgery, School of Medicine, Washington University in St. Louis, St. Louis, Missouri; Sever Institute, McKelvey School of Engineering, Washington University in St. Louis, St. Louis, Missouri; School of Medicine, Washington University in St. Louis, St. Louis, Missouri; Division of Cardiovascular Medicine, Department of Medicine, School of Medicine, Washington University in St. Louis, St. Louis, Missouri; Institute for Informatics, Division of General Medical Sciences, School of Medicine, Washington University in St. Louis, St. Louis, Missouri

## Abstract

**Background:** Early identification of heart failure patients at increased risk for near-term adverse outcomes would assist clinicians in efficient resource allocation and improved care. Deep learning can improve identification of these patients.

**Methods:** This retrospective study examined adult heart failure patients admitted to a tertiary care institution between January 2009 and December 2018. A deep learning model was constructed with a dense input layer, three long short-term memory (LSTM) layers, and a dense hidden layer to cohesively extract features from time-series and non-time-series EHR data. Primary outcomes were all-cause hospital readmission or death within 30 days after hospital discharge.

**Results:** Among a final subset of 49,675 heart failure patients, we identified 171,563 hospital admissions described by 330 million EHR data points. There were 22,111 (13%) admissions followed by adverse 30-day outcomes, including 19,122 readmissions (87%) and mortality in 3,330 patients (15%). Our final deep learning model achieved an area under the receiver-operator characteristic curve (AUC) of 0.613 and precision-recall (PR) AUC of 0.38.

**Conclusions:** This EHR-based deep learning model developed from a decade of heart failure care achieved marginal clinical accuracy in predicting very early hospital readmission or death despite previous accurate prediction of 1-year mortality in this large study cohort. These findings suggest that factors unavailable in standard EHR data play pivotal roles in influencing early hospital readmission.

**Clinical Perspective:** *What is new?:* We developed an EHR-based deep learning model trained by 330 million data points from one of the largest cohorts of heart failure patients to date. Despite this model’s highly accurate prediction of long-term outcomes, such as mortality and disease progression, our findings suggest that EHR data alone offers limited predictive power for predicting the short-term outcomes of 30-day hospital readmission or death.

*What are the clinical implications?:* Our study supports the notion that hospital readmission, in contrast to other health outcomes, is uniquely driven by additional factors beyond traditional EHR variables. Once identified, incorporation of these determinants into future deep learning models could allow for accurate heart failure risk-stratification at hospital discharge to facilitate more efficient allocation of limited resources to the most vulnerable patients.

## Introduction

Heart failure remains a significant cause of morbidity and mortality in the United States with an annual mortality rate ranging from 22-30% (1–5). Heart failure patients represent the most rapidly growing and resource-intensive subset among patients with cardiovascular disease (6,7). Additionally, patients with heart failure have among the highest rates of unplanned 30-day hospital readmissions, ranging from 20-56% (5–7), many of which may be preventable (5–11). To this end, there has been increased emphasis on identifying heart failure patients who are at increased risk for adverse outcomes early after hospital discharge.

Deep learning models employ automated learning techniques to identify outcome-predictive data patterns in large datasets (12,13). Prior investigations using machine learning models to predict hospital readmission in heart failure patients have reported inconsistent results (14–31). This may be attributed to difficulty in obtaining consistent, high-quality granular data for this heterogeneous patient group, or simply due to a lack of predictive signal in standard clinical metrics. Many other potential predictors, such as medication adherence and socioeconomic status, are not explicitly captured in EHR data but have been identified as critical determinants of care metrics including hospital readmission (32).

Although prior studies have not identified any single EHR variable capable of accurately predicting heart failure readmission with the precision required to drive clinical decision-making, predictive associations between groups of EHR variables and heart failure outcomes have been reported (17,24,32,33). As an alternative strategy to traditional regression analysis models, this investigation employs the pattern-recognition capabilities of deep learning to exploit the outcome-predictive capabilities associated with often complex and non-intuitive groupings of variables found in the medical records of heart failure patients.

EHR data platforms present patient-specific clinical variables in a uniquely structured format that may simplify and accelerate direct integration into machine learning models, facilitating application of their advanced analytic capabilities in risk prediction. These attributes give EHR-based deep learning models considerable potential for direct clinical utility in the identification of heart failure patients at high-risk for readmission or mortality. The accurate identification of these high-risk patients may improve distribution of available resources to the most vulnerable heart failure patients.

## Methods

### Data Source

The human studies *Institutional Review Board* at Washington University School of Medicine approved this study. All machine learning clinical variables analyzed in this study were obtained from the EHR of patients at Barnes-Jewish Hospital at Washington University Medical Center in St. Louis, Missouri.

### Cohort and Study Design

The study population consisted of adult patients between the ages of 18 to 90 years old with any heart failure International Classification of Diseases (ICD) codes admitted between January 2009 and December 2018. Admissions within 30 days before the conclusion of data collection or a length of stay (LOS) under 48 hours were excluded. Admissions with no discharge date or ending in death were excluded as *index* hospitalizations, although not as subsequent *readmissions*. Finally, at least 5 unstructured records (e.g., diagnosis, treatment, procedures, etc.), 1 vital sign record, and 1 laboratory value were required to include a recorded medical encounter as a hospitalization. These criteria helped exclude brief admissions scheduled for diagnostic purposes, such as cardiac catheterization and transesophageal echocardiography, which are often ordered on an outpatient basis but performed in the hospital due to their invasive nature.

The unit of analysis used to train our machine learning model was hospital admission, in contrast to other studies that have performed analysis at a patient (29) or unspecified (27) level. In patients with serial hospitalizations, each admission may qualify as a unique index hospitalization as well as a potential readmission if within 30 days after a prior discharge. By analyzing each instance as a unique opportunity for learning, data utilized for model training can be maximized. Moreover, this approach more accurately represents clinical practice and enhances prediction applicability, since heart failure clinicians are routinely tasked with evaluating the risk of readmission at the end of each hospitalization.

Our primary positive outcomes were all-cause readmission lasting ≥ 48 hours or death within 30 days after discharge following an index hospitalization. These were classified as “positive events”. The 30-day window after hospital discharge has been identified as a key health policy metric for health care quality improvement (34,35). Readmissions within this time frame often reflect poor patient risk evaluation and inadequate recognition of post-discharge needs (36). Ventricular assist device placement, extracorporeal membrane oxygenation (ECMO) support, or cardiac transplantation are not considered separately since all are captured as hospitalization events. Model inputs included EHR data prior to index hospitalization discharge date, including all EHR data obtained during prior and current hospitalizations and in the outpatient setting.

### Feature Extraction and Deep Learning Network Design

The deep learning model in **Figure 1** is comprised of three LSTM layers to extract features from time-series EHR data, including vital signs, medication, laboratory, assessment, procedural, and diagnostic coding variables (37). Additionally, a dense layer extracts non-time-series patient attributes, such as age, gender, race; and a dense hidden layer which extracts additional features from the output features generated by the three LSTM input layers and the dense input layer. The intermediate features are extracted and pooled to a final hidden dense layer containing 16 neurons and a bias neuron. The model employs the rectified linear activation unit function (ReLU) for all dense hidden layers, and sigmoid activation for the final output neuron.

**Figure 1.**
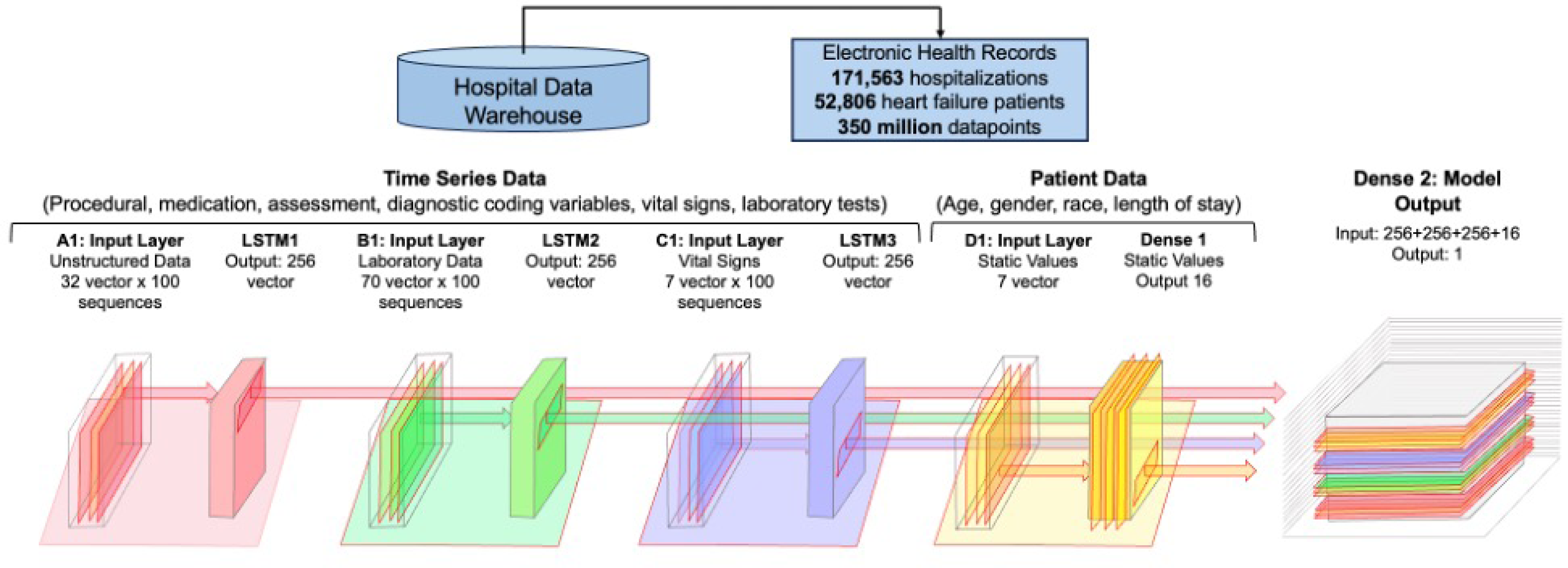
A deep learning model was constructed with a dense input layer, three long short-term memory (LSTM) layers, and a dense hidden layer to cohesively extract features from time-series and non-time-series EHR data.

Most patients had available data for a subset of the 70 laboratory values evaluated (**Table 1**). These values are encoded by a 70-length vector which initially starts as the mean data set value for each laboratory test. As new results are obtained over time, the previous value in the vector is replaced and remains in place until new testing is performed. Therefore, the vector contains the most recent test results or the mean value for the dataset if no results are available for a specific patient. Vectors are entered as input sequences for the LSTM layers in a sliding window fashion with the oldest vectors sliding off once they reach the end of the sequence. Vital signs are similarly encoded using a 7-length vector whose values are updated as more recent vital signs are recorded in the EHR.

**Table 1.** Univariate importance of the 70 clinical variables evaluated for individual impact on 30-day all-cause readmission and mortality.

| Variable | AUC Without Variable | Importance |
| --- | --- | --- |
| **Vital Signs** |  |  |
| Height | 0.593 | 1.000 |
| Weight | 0.594 | 0.996 |
| Heart rate | 0.598 | 0.987 |
| Diastolic blood pressure | 0.598 | 0.987 |
| Oxygen flow rate | 0.599 | 0.984 |
| Systolic blood pressure | 0.603 | 0.974 |
| Oxygen saturation | 0.603 | 0.974 |
| **Laboratory Values** |  |  |
| C-reactive protein | 0.503 | 1.000 |
| Sodium | 0.510 | 0.987 |
| Lactate dehydrogenase | 0.513 | 0.980 |
| Brain natriuretic peptide | 0.517 | 0.973 |
| Hematocrit | 0.521 | 0.966 |
| Fasting glucose | 0.521 | 0.965 |
| Monocytes | 0.524 | 0.958 |
| Albumin | 0.525 | 0.956 |
| Fraction of inspired oxygen | 0.526 | 0.954 |
| Lactic acid | 0.529 | 0.949 |
| Uric acid | 0.529 | 0.948 |
| Plasma potassium | 0.530 | 0.946 |
| Ammonia | 0.531 | 0.945 |
| Cannabinoids | 0.532 | 0.943 |
| Hemoglobin | 0.533 | 0.940 |
| Amylase | 0.534 | 0.939 |
| Total hemoglobin | 0.535 | 0.936 |
| Venous partial pressure of oxygen | 0.537 | 0.932 |
| Triglycerides | 0.541 | 0.925 |
| Hemoglobin A1c | 0.541 | 0.925 |
| Chloride | 0.541 | 0.925 |
| Potassium | 0.541 | 0.924 |
| Carbon dioxide | 0.545 | 0.916 |
| Carboxyhemoglobin | 0.546 | 0.915 |
| Calculated oxygen saturation | 0.546 | 0.914 |
| Venous oxygen content | 0.546 | 0.914 |
| Oxygen gradient | 0.547 | 0.912 |
| Ketones | 0.550 | 0.907 |
| Alanine aminotransferase | 0.550 | 0.906 |
| Basophils | 0.552 | 0.903 |
| Base excess | 0.552 | 0.901 |
| Methemoglobin | 0.553 | 0.901 |
| Blood urea nitrogen | 0.553 | 0.899 |
| Troponin I | 0.555 | 0.897 |
| Benzodiazepines | 0.556 | 0.894 |
| Pulmonary artery oxygen | 0.558 | 0.891 |
| Partial thromboplastin time | 0.558 | 0.891 |
| Magnesium | 0.558 | 0.890 |
| Alkaline phosphatase | 0.559 | 0.888 |
| Glucose | 0.560 | 0.887 |
| Partial pressure of carbon dioxide | 0.561 | 0.885 |
| Pulmonary artery oxyhemoglobin | 0.563 | 0.880 |
| International normalized ratio | 0.563 | 0.879 |
| Cocaine | 0.564 | 0.878 |
| Creatinine | 0.569 | 0.867 |
| Direct bilirubin | 0.572 | 0.862 |
| High-density lipoprotein | 0.574 | 0.858 |
| Prothrombin time | 0.575 | 0.856 |
| Phosphorous | 0.575 | 0.855 |
| D dimer | 0.584 | 0.839 |
| Aspartate aminotransferase | 0.586 | 0.833 |
| Fraction of inspired oxygen | 0.595 | 0.816 |
| Plasma protein | 0.595 | 0.815 |
| Anion gap | 0.596 | 0.813 |
| Calcium | 0.598 | 0.809 |
| pH | 0.599 | 0.807 |
| Eosinophils | 0.599 | 0.807 |
| Fibrinogen | 0.600 | 0.806 |
| Urine specific gravity | 0.600 | 0.806 |
| Lymphocytes | 0.600 | 0.806 |
| Indirect Coombs | 0.600 | 0.806 |
| Lipase | 0.600 | 0.805 |
| Bicarbonate | 0.602 | 0.802 |
| Creatine kinase | 0.602 | 0.802 |
| Platelets | 0.602 | 0.802 |
| Bilirubin | 0.603 | 0.800 |
| Partial pressure of oxygen | 0.603 | 0.800 |
| Thyroid stimulating hormone | 0.603 | 0.799 |
| Neutrophils | 0.603 | 0.799 |
| Cholesterol | 0.604 | 0.798 |
| Oxygen saturation | 0.604 | 0.798 |
| **Static Variables** |  |  |
| Unstructured row count | 0.540 | 1.000 |
| Laboratory test row count | 0.567 | 0.940 |
| Length of stay (LOS) hours | 0.591 | 0.889 |
| Vital sign row count | 0.597 | 0.875 |
| Gender | 0.602 | 0.865 |
| Race | 0.604 | 0.859 |
| Age at admission | 0.606 | 0.857 |

The coding variables for basic procedures, surgical procedures, diagnostics, assessments, and medications were transformed into 100-length sequences of 32-dimensional vectors using Python’s Genism Word2Vec (38). These sequences were then entered into an LSTM layer. Since the data were high-cardinality and sparse, no missing data management was required. The encoding process only considered present values and ignored those that were not present in the time series.

The data were randomly divided into three sets: training (80%), validation (10%), and testing (10%). The model’s performance was evaluated using AUC, PR AUC, log loss, and Brier score. Feature importance was determined using the permutation feature importance method, where each feature in the trained neural network was permuted and the change in AUC analyzed. Additionally, multiple features were permuted together to analyze grouped feature importance.

## Results

### Heart Failure Dataset Baseline Characteristics

Our initial patient cohort comprised 52,806 heart failure patients. After the previously described exclusions, our final study set consisted of 49,675 patients, of whom, 20% (9,975) experienced positive events during the 10-year study period. Of those who experienced positive events, 84% (8,419) experienced 30-day readmissions and 28% (2,751) suffered 30-day all-cause mortality. The baseline demographic characteristics and vital signs of our training and internal validations cohorts are listed in **Table 2**.

**Table 2.**
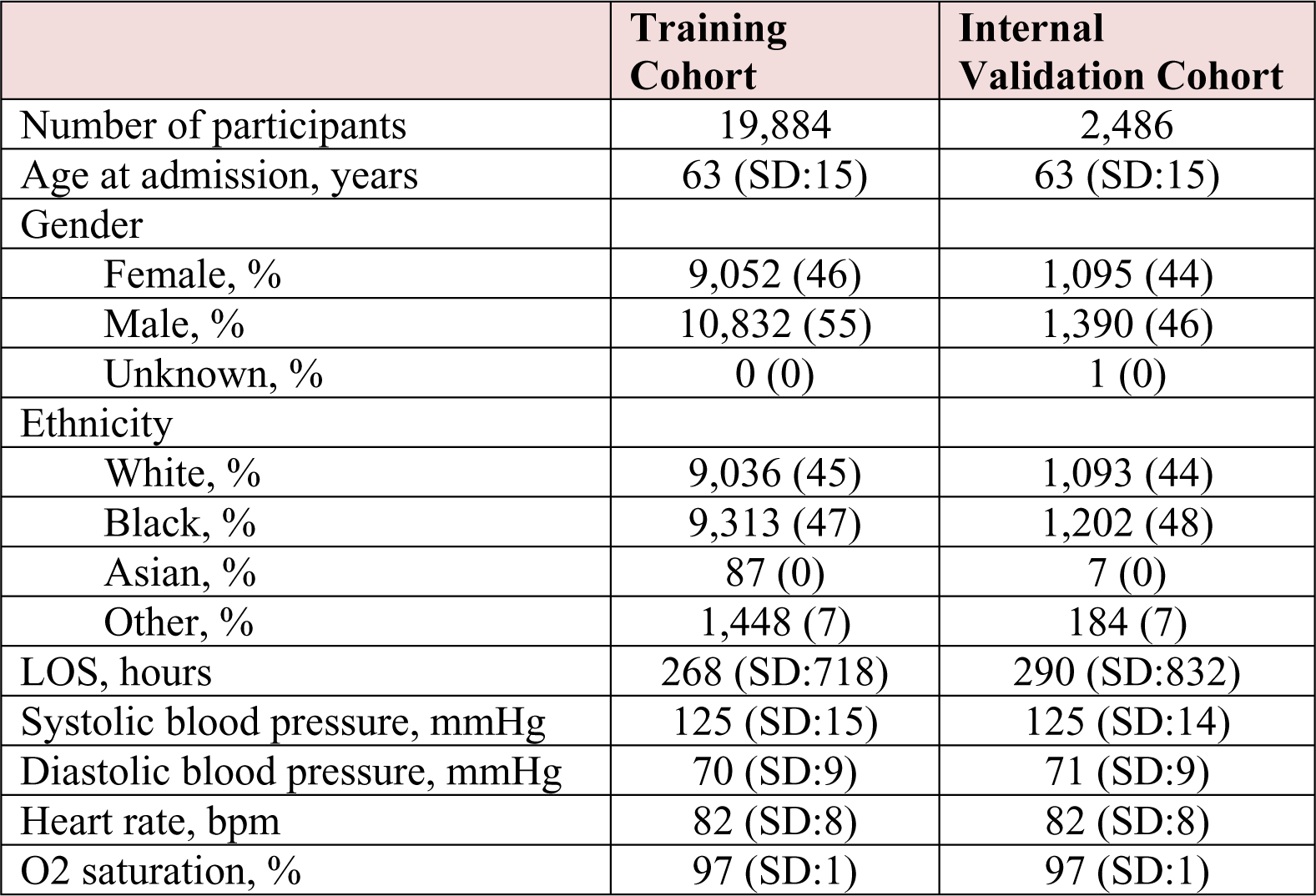
Baseline characteristics in training and internal validation cohorts.

|  | **Training Cohort** | **Internal Validation Cohort** |
| --- | --- | --- |
| Number of participants | 19,884 | 2,486 |
| Age at admission, years | 63 (SD:15) | 63 (SD:15) |
| Gender |  |  |
| Female, % | 9,052 (46) | 1,095 (44) |
| Male, % | 10,832 (55) | 1,390 (46) |
| Unknown, % | 0 (0) | 1 (0) |
| Ethnicity |  |  |
| White, % | 9,036 (45) | 1,093 (44) |
| Black, % | 9,313 (47) | 1,202 (48) |
| Asian, % | 87 (0) | 7 (0) |
| Other, % | 1,448 (7) | 184 (7) |
| LOS, hours | 268 (SD:718) | 290 (SD:832) |
| Systolic blood pressure, mmHg | 125 (SD:15) | 125 (SD:14) |
| Diastolic blood pressure, mmHg | 70 (SD:9) | 71 (SD:9) |
| Heart rate, bpm | 82 (SD:8) | 82 (SD:8) |
| O2 saturation, % | 97 (SD:1) | 97 (SD:1) |

### Heart Failure Dataset 30-day Adverse Events

Of 171,563 study hospitalizations, 22,111 (13%) met our primary positive outcome (**Table 3**). Of these positive events, 19,122 (11%) involved 30-day all-cause readmission and 3,330 (2%) involved 30-day all-cause mortality. Some patients experienced both mortality and re-admission within 30 days (**Table 3**).

**Table 3.** Comparison of positive and negative observations.

|  | **Negative**<br>**Observation** | **Positive**<br>**Observation** |
| --- | --- | --- |
| Number of participants | 49,675 | 9,975 |
| Age at admission, years | 62 (SD:13) | 62 (SD:15) |
| Gender |  |  |
| Female, % | 22,956 (46) | 4,488 (45) |
| Male, % | 26,717 (54) | 5,487 (55) |
| Unknown, % | 2 (0) | 0 (0) |
| Ethnicity |  |  |
| White, % | 19,373 (39) | 3,591 (36) |
| Black, % | 27,321 (55) | 5,386 (54) |
| Asian, % | 496 (1) | 1 (0) |
| Other, % | 2,485 (5) | 997 (10) |
| LOS, hours | 98 (SD:453) | 128 (SD:439) |
| Systolic blood pressure, mmHg | 127 (SD:20) | 125 (SD:21) |
| Diastolic blood pressure, mmHg | 71 (SD:13) | 71 (SD:13) |
| Heart rate, bpm | 81 (SD:13) | 82 (SD:13) |
| O2 saturation, % | 97 (SD:2) | 97 (SD:2) |

### Heart Failure Dataset Hospital LOS Data

The overall average LOS for the index hospitalizations was 122 hours. The average LOS of positive versus negative observations was 148 hours versus 133 hours, respectively.

### Heart Failure Dataset Variable Importance

**Table 1** shows the importance of the 70 variables evaluated for individual impact on 30-day all-cause readmission and mortality. Patient height, C-reactive protein level, and unstructured row count (which included variables such as diagnosis, treatment, procedures, etc.) demonstrated the highest importance at a univariate level. **Figure 2** demonstrates the relative predictive importance of vital sign and laboratory test variables depicted on the x and y axes, respectively. Variables are listed in descending order of univariate importance. Red hues represent those with highest impact on model prediction (darker red hues representing factors with the most important impact) and blue hues highlighting low impact variables (dark blue hues representing the least important impact). Brain natriuretic peptide levels and serum sodium levels exhibited the highest impact among the clinical variables evaluated (vertical gradient), and there was no apparent association between clinical variable importance and vital sign values (horizontal gradient).

**Figure 2.**
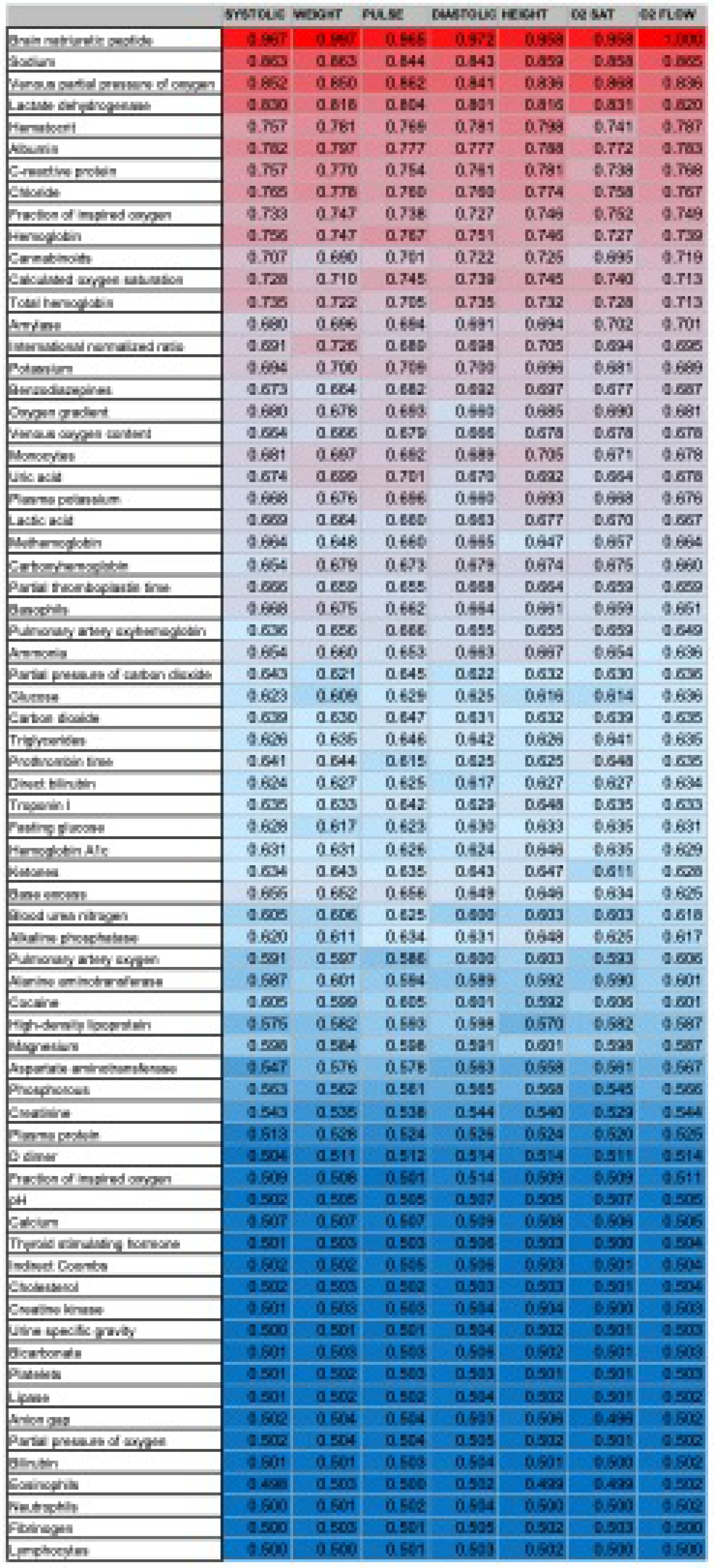
Heat map demonstrating the relative predictive importance of vital sign and laboratory test variables depicted on the x and y axes, respectively. Variables are listed in descending order of univariate importance, with red hues representing those with highest impact on model prediction and blue hues highlighting low impact variables.

### Model Performance

The performance of the model for predicting 30-day readmission or death in heart failure patients was measured using the ROC curve with a test set ROC AUC of 0.613 (95% CI: 0.59***-*** 0.64) (**Figure 3**). **Table 4** shows additional performance metrics, including PR AUC of 0.38 (CI: 0.33*-*0.41) shown in **Figure 4**. The model’s predictive performance remained stable across a range of hospitalization durations, including hospitalizations with a brief LOS (**Table 1**). The log loss was 0.57 with a Brier Score of 0.19

**Figure 3.**
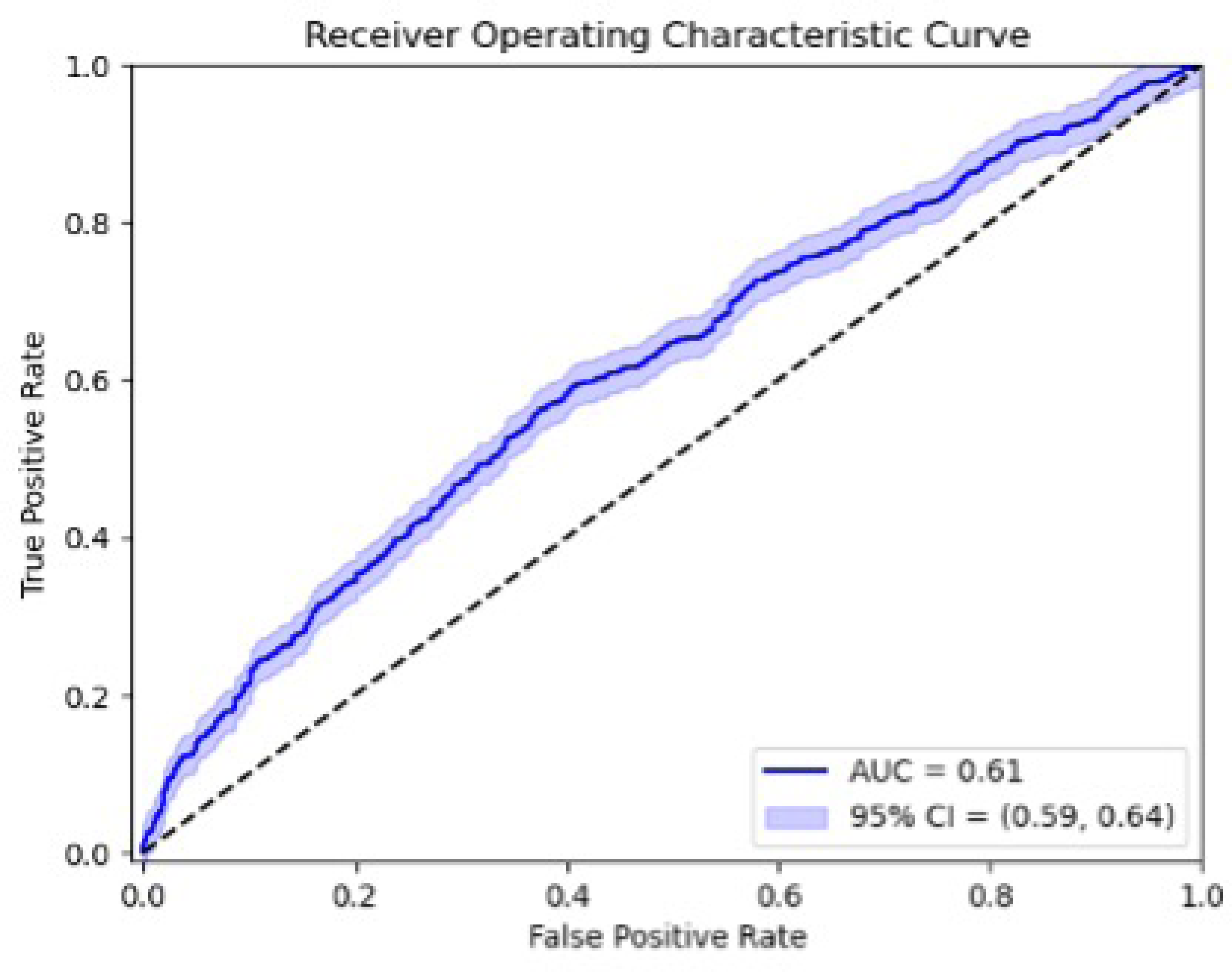
The area under the receiver-operating characteristic curve (AUC) (or C-statistic) of 0.61 with a 95% CI of 0.59-0.64 demonstrates marginal clinical accuracy in short-term prediction of adverse 30-day outcomes in this large study cohort.

**Table 4.** Predictive model performance results.

| Measure | Result |
| --- | --- |
| ROC AUC | 0.613 |
| Log Loss | 0.570 |
| PR AUC | 0.380 |
| Brier Score | 0.190 |

**Figure 4.**
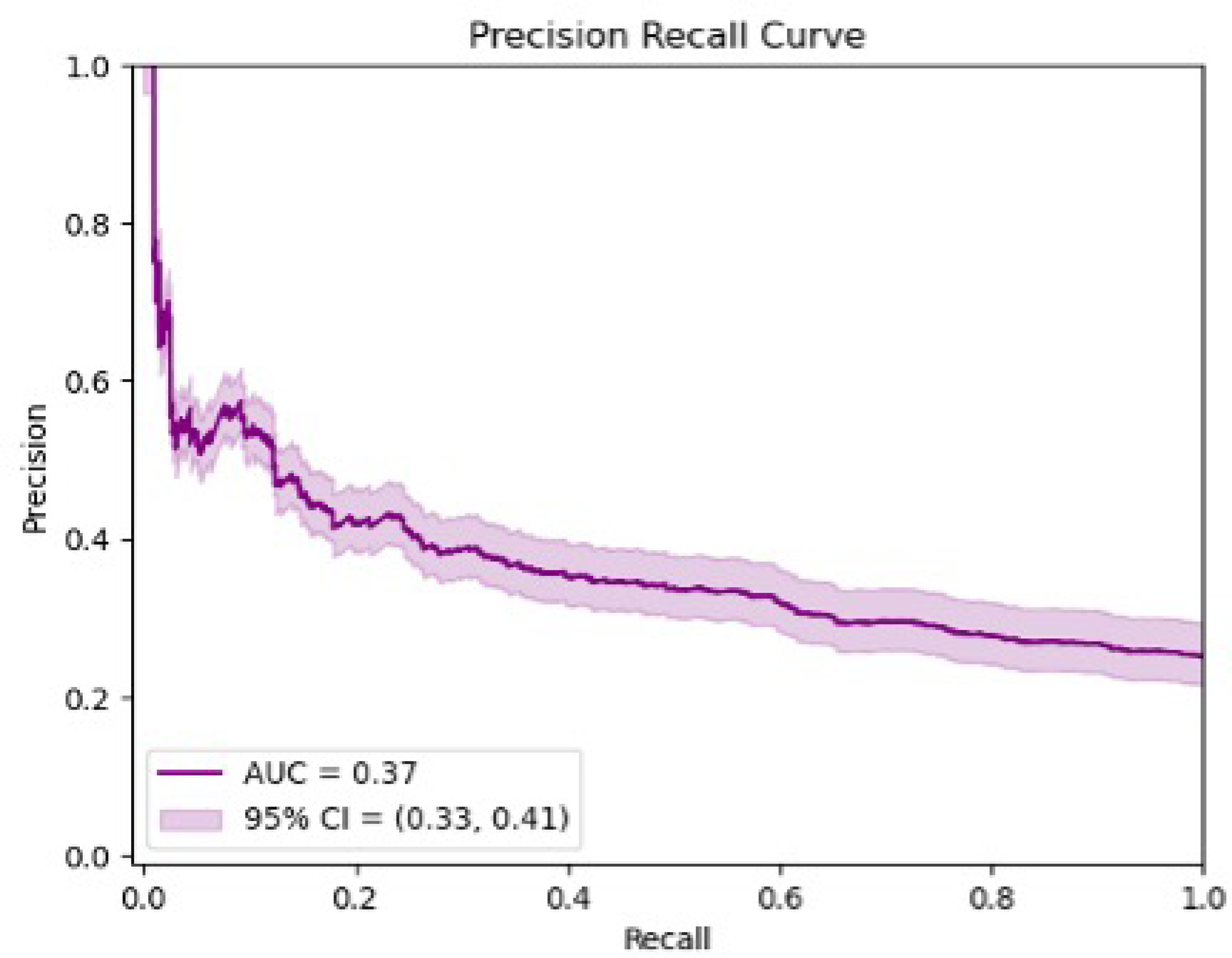
The precision recall area under the curve (PR AUC) of 0.37 with a 95% CI of 0.33-0.41 further supports the notion that standard EHR data lacks crucial factors, such as socioeconomic determinants, that lay pivotal roles in influencing early readmission and mortality in this cohort.

## Discussion

This deep learning model predicting 30-day all-cause hospital readmission or death was developed using a decade of EHR data from a tertiary care hospital and included 330 million data points from 171,563 hospital admissions in 49,675 heart failure patients. The model employed time-sensitive LSTM networks to analyze subtle fluctuations in clinical EHR variables over time and utilized densely connected networks for fixed, non-time-dependent demographics. Notably, we previously demonstrated that a nearly identical model successfully generated accurate predictions of long-term adverse outcomes in the same cohort of heart failure patients, achieving an AUC of 0.91 (39). Our current findings indicate that EHR data alone is insufficient to generate similar predictive accuracy of 30-day hospital readmission or death in heart failure patients. This study underscores the unique nature of hospital readmission in comparison to other health quality metrics and the need to identify additional factors beyond traditional EHR variables that influence preventable hospital readmissions in this patient population.

Our study supports a growing body of literature heightening awareness of socioeconomic factors (such as income, education, place of residence, disability, ethnicity, race) as pivotal determinants contributing to hospital readmission in heart failure patients (40,41). While it is unlikely that targeting individual socioeconomic factors alone will be enough to reduce readmissions (41), the predictive model we seek likely requires cohesive integration of a cumulative burden of socioeconomic determinants in combination with key clinical variables that can be extracted from the EHR. This task is one for which machine learning is uniquely suited, given its unique proficiency at identifying non-intuitive, multiparametric outcome predictive patterns, and its seemingly boundless capability for comprehensive data analysis and integration (14,15,17,19,22–24). Additionally, incorporating time-based metrics through recurrent neural network units allows for the detection of even subtle progression in heart failure status, which can uncover time-dependent relationships between variables to enhance prediction and accelerate learning. These results suggest only modest capability of time-sensitive neural networks for predicting post-discharge adverse outcomes within the framework of each individual patient’s unique heart failure course.

Prior research utilizing machine learning to predict hospital readmission in heart failure patients has yielded varying degrees of accuracy, with predictive AUC values ranging from 0.62 to 0.96 (14,15,17,19,22–24,26–29). Our final deep learning model was developed in one of the largest cohorts of heart failure patients to date and lends support to previous model results at the lower end of that spectrum of predictive accuracy. Although our model AUC does not attain the high level of accuracy achieved by Lv et al in a smaller cohort of 13,600 patients (17), our results are comparable to those reported by larger models and may accurately represent current potential of large scale implementation of patient-specific outcome prediction using an embedded EHR model.

By excluding hospitalizations with LOS ≤ 48 hours as index admissions or readmissions, our model attempts to exclude routine follow-up admissions, such as those related to elective, regularly scheduled endomyocardial biopsy and other surveillance investigations, many of which are unique to this complex patient subpopulation. This type of readmission may have contributed to overestimation of readmission for actual heart failure decompensation in previously published investigations (14,24,28). Instead, our model focuses upon longer hospitalizations, such as those for severe heart failure decompensation.

Several limitations must be considered when interpreting the findings of this study. Its retrospective design and the use of data from a single institution may restrict the generalizability of the model. Additionally, retrograde leakage of data that may falsely improve predictive accuracy is always a possibility (42–44). Our definition of a positive event as all-cause readmission or mortality may thereby include a small proportion of patients for whom 30-day readmission or death was not related to heart failure. Although not ideal, this primary outcome definition is the most effective and clinically applicable in our large population of medically complex heart failure patients. The inclusion of multiple hospitalizations from each patient in predictive model development may potentially skew results to the subpopulation with heightened disease severity and worse overall health status. However, multiple readmissions are common in heart failure patients and their impact should be represented in predictive models. Clinical models assist but should not substitute clinical judgement. Additional validation is essential to verify the applicability of this model in other heart failure patient populations.

Our results suggest that EHR data alone offers limited predictive power for certain care quality metrics, such as early hospital readmission or death, and support the notion that other factors not routinely recorded in hospital medical records may play a pivotal role in determining these outcomes. To this end, further investigation is necessary to determine which of these non-standard metrics are most important in this patient population. Once identified, incorporation of these variables into future deep learning models could allow for accurate heart failure risk-stratification at hospital discharge to facilitate more efficient allocation of limited resources to the most vulnerable patients.

## Data Availability

Data is available upon request. Provision of patient-protected data is not possible to protect patient identity.

## Abbreviations

AUC: Area under the receiver operating characteristic curve
BMI: Body mass index
ECMO: Extracorporeal membrane oxygenation
EHR: Electronic health record
ICD: International Classification of Diseases
LOS: Length of stay
LSTM: Long short-term memory
PR AUC: Precision-recall area under the curve
ReLU: Rectified linear activation unit function

